# The role of blame-related action tendencies in the vulnerability to major depressive disorder

**DOI:** 10.1101/2020.11.15.20232058

**Authors:** Suqian Duan, Andrew Lawrence, Lucia Valmaggia, Jorge Moll, Roland Zahn

## Abstract

**Background:** Persisting self-blaming emotional biases were previously associated with vulnerability to major depressive disorder (MDD). More specifically self-contempt/disgust biases distinguished remitted MDD, compared with never-depressed control participants. The contribution of action tendencies to MDD vulnerability and their relationship with blame-related emotions to prepare for subsequent behaviour is elusive. Here, we investigated whether maladaptive action tendencies such as “creating a distance from oneself” and “hiding” are associated with MDD vulnerability, as well as with self-disgust/contempt and shame respectively.

**Methods:** 76 participants with medication-free remitted MDD and 44 healthy control (HC) participants without a personal or family history of MDD completed the value-related moral sentiment task, which measured their blame-related emotions during hypothetical social interactions and a novel task to assess their blame-related action tendencies.

**Results:** As predicted, the MDD group exhibited a higher proneness to feeling like hiding and creating a distance from themselves compared with the HC group. Interestingly, apologising for one’s wrongdoing, was associated with all self-blaming emotions including shame, guilt, self-contempt/disgust and self-indignation, but was more common in HC. In contrast, apologising and perceiving to be in control of one’s friend’s wrongdoings were more common in MDD. Although shame was indeed associated with hiding, this was also true of guilt. Self-disgust/contempt was associated with attacking rather than creating a distance from oneself.

**Conclusions:** MDD vulnerability was associated with specific maladaptive action tendencies which were not clearly predicted by the type of emotion, thus unveiling novel cognitive markers and neurocognitive treatment targets.

**General Scientific summary:** This study confirmed the hypothesis that specific maladaptive action tendencies related to self-blame, such as feeling like hiding and feeling like creating a distance from oneself, were distinctive of people with major depressive disorder, even on remission of symptoms. These action tendencies were not clearly predicted by the type of emotion experienced, showing the importance of assessing them directly. This calls for novel psychological and neurocognitive treatments specifically aiming at maladaptive action tendencies which have so far not been directly addressed in standard assessments and treatments.

## Introduction

Previous studies have demonstrated the significance of moral emotions and self-blaming emotional biases as vulnerability factors for major depressive disorder (MDD; (Janoff-Bulman, Sheikh, & Hepp, 2009; O’Connor, Berry, Weiss, & Gilbert, 2002; Tangney, Stuewig, & Mashek, 2007; Zahn, Lythe, Gethin, Green, Deakin, Young, et al., 2015). Self-blaming emotions were hypothesized to be associated with action tendencies (Haidt, 2003; Janoff-Bulman et al., 2009; Tangney et al., 2007) which describe a motivational and cognitive state in which there is an increased tendency to engage in certain goal-related behaviours (Haidt, 2003), such as “creating a distance from someone”. Determining whether these action tendencies play a role in the vulnerability to MDD is an essential step not only towards a better understanding of the psychopathology, but also towards designing novel interventions and risk prediction markers.

As proposed by the revised learned helplessness model, one central feature of cognitive vulnerability to MDD is a tendency to excessively blame oneself for negative events occurring in life (Abramson, Seligman, & Teasdale, 1978). In line with the theory, imbalances in blame-related emotions have been shown to be closely related to individuals’ negative mental health outcomes and risks of MDD. For example, overgeneralised guilt (O’Connor et al., 2002) and shame have been observed in people with MDD even on remission (Green, Moll, Deakin, Hulleman, & Zahn, 2013) and associated with depressive symptoms in those without a clinical diagnosis of MDD (Tangney et al., 2007). In a further study (Zahn, Lythe, Gethin, Green, Deakin, Workman, et al., 2015), individuals with remitted MDD exhibited a self-contempt/disgust bias and a reduction in contempt/disgust towards others. As the risk of depressive episodes is increased in people with MDD even after a single episode, rising from around 15% to 50% (Eaton et al., 2008), understanding the differences between remitted MDD and healthy control groups is a first step to identify potential vulnerability traits associated with MDD (Bhagwagar & Cowen, 2008). Thus, previous findings demonstrated the potential role of self-blaming emotions as vulnerability traits for MDD that remain present during remission.

Despite the importance of previous findings, existing measures of self-blaming emotional biases have some critical limitations, one of which is that their mechanism in motivating adaptive or maladaptive social actions is elusive. As proposed by Tangney et al. (2007), adaptive moral emotions promote constructive and proactive pursuit, whereas maladaptive moral emotions motivate defensiveness, social withdrawal and interpersonal separation (Tangney et al., 2007). In addition, this difference is possibly determined by an individuals’ action tendencies associated with their moral emotions (Haidt, 2003; Tangney et al., 2007). Action tendencies are fundamental characteristics of emotions that closely relate to emotion differentiation and its evolutionary function for the social survival of individuals (Roseman, Wiest, & Swartz, 1994). Previous researchers have proposed associations between moral emotions and action tendencies as well as the adaptive or maladaptive nature of their relationships. For example, Tangney and colleagues conceived of guilt as being more strongly associated with adaptive action tendencies such as reparative actions including confessions and apologies, whereas they operationalised shame as associated with maladaptive action tendencies such as an attempt to deny, hide, or escape the shame-inducing situation (Haidt, 2003; Janoff-Bulman et al., 2009; Tangney et al., 2007). However, other researchers argued that both guilt and shame can be adaptive or maladaptive depending on the specific circumstances, and maladaptive and overgeneralised forms of guilt (O’Connor, Berry, Weiss, Bush, & Sampson, 1997) were increased in current (O’Connor et al., 2002) and remitted MDD (Karen E. Lythe et al., 2020).

Indeed, the nature of action tendencies (adaptive vs. maladaptive) might be important in guiding the behavioural consequences of moral emotions, thereby providing a direct link to vulnerability to psychopathology (O’Connor et al., 1997; Tangney et al., 2007). Of particular relevance in MDD is the action tendency associated with contempt/disgust and shame, as self-directed contempt/disgust biases were associated with MDD vulnerability (Zahn, Lythe, Gethin, Green, Deakin, Workman, et al., 2015) and shame proneness was associated with higher depressive symptoms in both clinical and non-clinical populations (Tangney et al., 2007). Both self-contempt/disgust and shame entail the devaluation of one’s character (Fischer & Roseman, 2007), but the former is more strongly related to violations of internalised moral values (Higgins, 1987). The feeling of contempt/disgust has mostly been investigated when directed towards others and plays a role in separating a group or individual from others that are thought to be below one’s own social status or against one’s moral standard (Haidt, 2003). Thus, contempt/disgust towards others was proposed to be associated with a motivation to avoid, expel or withdraw from an offensive object or person (Haidt, 2003; Rozin, Haidt, & McCauley, 1993). In empirical investigations, contempt and disgust towards others consistently predicted an action tendency to move away from an antagonistic social group (Mackie & Smith, 1998). We hypothesised that self-contempt/disgust would be associated with a tendency to create a distance from oneself, analogous to the literature on contempt/disgust directed at others. In addition, shame was found to trigger postural signs of defence and self-concealment, which motivates people to deny, hide or escape the shame-inducing situations (Dickerson, Gruenewald, & Kemeny, 2004; Tangney et al., 2007). These evidence pointed to the potential importance of feeling like creating a distance from oneself and hiding, which are both associated with avoiding rather than fixing difficult social interactions, in the vulnerability to major depressive disorder, where social withdrawal is one of the most consistent symptoms (Zahn, Lythe, Gethin, Green, Deakin, Young, et al., 2015).

Despite the potential distinctive psychopathology of feeling like creating a distance from oneself and feeling like hiding, to our knowledge, these action tendencies have so far not been directly assessed. Furthermore, there is no systematic investigation of how action tendencies and moral emotions are linked in MDD. The present study aimed to elucidate these questions by directly examining the relationship between blame-related emotions and action tendencies, and their potential role in MDD vulnerability. We developed a novel action tendency task that specifically assessed different action tendencies (feeling like: apologising, hiding, creating a distance from oneself, attacking oneself) when people experienced self-blame-related emotions (shame, guilt, self-contempt/disgust, self-directed anger) and comparing them against emotions and action tendencies associated with blaming others. We hypothesised that individuals with medication-free and fully remitted MDD were more likely to have maladaptive action tendencies reflecting their vulnerability to further episodes despite current symptom remission, when compared with a control group without a personal and family history of MDD. According to Tangney et al. (2007), maladaptive action tendencies include those that trigger interpersonal separation and motivate people to deny or hide the self-blame-inducing situations. More specifically, based on our previous work, we firstly hypothesised that individuals with MDD have an increased tendency to create a distance from themselves and/or to hide as described above. Secondly, we hypothesized that self-contempt/disgust is associated with feeling like creating a distance from oneself and thirdly, that shame is distinctively associated with feeling like hiding.

## Methods

### Participants

Seventy-six medication-free participants with remitted MDD and 44 healthy control (HC) participants took part in the study and completed both value-related moral sentiment (VMST) and action tendency task. Participants were recruited via online and print advertisements as part of the project: Development of Cognitive and Imaging Biomarker Predicting Risk of Self-Blaming Bias and Recurrence in Major Depression” funded by the UK Medical Research Council and results of the VMST and psychopathological characteristics have been previously reported (Zahn, Lythe, Gethin, Green, Deakin, Workman, et al., 2015; Zahn, Lythe, Gethin, Green, Deakin, Young, et al., 2015). A total of 707 people took part in an initial phone screening interview to establish whether they would be invited to a clinical assessment using the Structured Clinical Interview-I for DSM-IV (First, Gibbon, Spitzer, Benjamin, & Williams, 1997). The main inclusion criteria were a diagnosis of MDD and a remission period for at least six months for the MDD group, and no history of an axis-I disorder or first-degree relatives with mood disorders or schizophrenia for the HC group.

After the initial phone screening interview, 276 people passed the screening with 184 in the MDD group and 92 in the HC group (431 people were excluded at this stage, the exclusion reasons following the phone screening interview are listed in Supplementary Table 1). Participants were then invited to see a senior psychiatrist (RZ) and take part in a face-to-face clinical assessment to further exclude the possibility of current co-morbid axis-I and relevant past axis-I disorders (full inclusion and exclusion criteria and assessment details can be found in Zahn, Lythe, Gethin, Green, Deakin, Workman, et al. (2015). Following the face-to-face assessment, 96 participants with MDD and 48 HC participants met all the criteria and took part in the current study. Twenty participants with MDD and 4 HC participants were excluded because they only completed the value-related moral sentiment task (VMST) and did not complete the action tendency task. This left data from 76 participants with MDD and 44 HC participants to be included in the final analysis. Demographic information and clinical characteristics of the included participants is shown in Table 1. There were no significant differences regarding the age, sex, nor the years of education of the two groups. As to be expected, depressive symptoms of MDD participants were slightly but significantly higher than those of HC participants as measured by the Beck Depression Inventory (Beck, Ward, Mendelson, Mock, & Erbaugh, 1961) total score and the Montgomery-Asberg Depression Rating Scale (Montgomery & Åsberg, 1977) total score.

**Table 1.** Demographic characteristics of participants.

|  | **Sample** |  | **p-value** |
| --- | --- | --- | --- |
|  | **rMDD**<br>**(n=76)** | **HC**<br>**(n=44)** |  |
| Age | 35.9 (13.1) | 33.7 (12.9) | 0.379 |
| Sex*[Female] | 55 (72.4%) | 28 (63.6%) | 0.428 |
| Years of Education | 16.8 (2.16) | 17.3 (2.48) | 0.245 |
| BDI Score | 3.71 (3.73) | 0.84 (1.63) | <0.001 |
| MADRS Score | 1.08 (1.49) | 0.59 (1.19) | 0.051 |
| GAF Score | 85.3 (5.87) | 89.1 (2.60) | <0.001 |

BDI = Beck Depression Inventory; MADRS = Montgomery-Asberg Depression Rating Scale; GAF - Global Assessment of Function. Values are Mean (Standard Deviation) for approximately continuous variables and Count (Percentage%) for categorical variables. Summary p-values are obtained from Welch's t-tests and Fisher's exact test respectively.

### Ethical approval

This study was approved by the South Manchester NHS Research Ethics Committee. All participants have given written informed consent after the procedures of the study have been fully explained.

### Assessment of blaming emotions and action tendencies

After assessing blame-related emotions using the value-related moral sentiment (VMST), which has been described and validated in our previous studies (Green, Lambon Ralph, Moll, Deakin, & Zahn, 2012; K. E. Lythe et al., 2015), participants were asked to complete the novel action tendencies task at home using excel macros, following their baseline assessment. The task was developed by our research team for the purpose of this study. At the beginning of the VMST, participants were asked to enter the name of their best friend. Then they were presented with sentences containing hypothetical social interactions in which either the participant (in the self-agency condition) or their best friend (in the other-agency condition) acts contrary to social and moral values. The same social interactions were used for both self-agency and other-agency conditions, with 90 trials in each condition. 50% of the trials used negative social behaviours (e.g., does act stingily) and 50% used negated positive social behaviours (e.g., does not act generously). In the VMST, participants were asked to rate the unpleasantness of each social interaction using a 1-7 point Likert scale, where 1 indicates not unpleasant at all and 7 indicates extremely unpleasant. They were also required to choose the feeling that they would feel most strongly from different self-blaming emotions (shame, guilt, contempt/disgust towards self and indignation/anger towards self), other-blaming emotions (contempt/disgust towards friend or indignation/anger towards friend) or no/other emotions. Self-blaming and other-blaming trials were defined as those that were perceived as highly unpleasant (those rated at the individual median or above in the VMST) in both self-agency and other-agency conditions.

In the novel action tendency task, all the hypothetical social interactions were shown again. Participants were instructed to select one action that they would most strongly feel like doing from eight different action tendencies (feel like verbally or physically attacking/punishing your friend, feel like verbally attacking or physically attacking/punishing yourself, feel like apologizing/fixing what you have done, feel like hiding, feel like creating distance from your best friend, feel like creating distance from yourself, no action, other action). Participants were also asked to rate how responsible they would feel, and how much control they felt they would have for each social action using a 1-7 point Likert scale, where 1 indicated “not at all” and 7 indicated “extremely/completely”.

### Data analysis

All statistical analyses in the study were carried out using R software. A complete case approach was taken for each planned analysis To test our first hypothesis (individuals with MDD had an increased tendency to create a distance towards themselves and/or hide), a repeated measures ANOVA was conducted to examine whether the proportion of trials selected by participants differed by action tendency, agency condition (self-vs. other) and clinical group and whether there were interactions between the three variables. Post-hoc tests for between-group differences in each action tendency over both agencies were conducted. Multiple comparison corrections were carried out for all post-hoc tests.

To test our second and third hypotheses (self-contempt/disgust was associated with feeling like creating a distance from oneself and shame was associated with feeling like hiding), the relationships between moral emotions and each action tendency were tested. Using R package lme4, separate mixed effect logistic regression models were conducted for each action tendency as outcome variable, with its agency-congruent moral emotions, group (MDD vs. HC) as well as their interactions as predictors. Trials in the VMST and action tendency task were included if only one moral emotion and one action tendency were selected. Participants were excluded if they chose more than one moral emotion or action tendency in more than 5% of the trials (9 trials). This ensured that all participants included in the logistic regression models understood the instruction of the tasks and could distinguish different moral emotions and action tendencies. In addition, data were only included if participants perceived a trial as highly unpleasant (those rated at the individual median or above) in the respective self-agency and other-agency conditions. Reference categories for moral emotions in all models were the trials in which participants chose no/other emotion. The significance threshold was set to an approximate Bonferroni-corrected p<.05 across all 6 models, corresponding to an uncorrected p <.008 in each model.

Perception of control in the hypothetical social scenarios were compared between two groups using Welch’s t-tests. In addition, to examine the relationship between clinical variables and action tendencies, Kendall’s rank correlations were used to test the correlation between proportions of choosing each maladaptive action tendency, number of previous depressive episodes, and participants’ BDI scores.

Agency-congruent moral emotions included all self-blaming emotions (shame, guilt, self-disgust/contempt and self-indignation) for self-agency-related action tendencies: feeling like apologising, hiding, creating a distance from oneself, or attacking oneself and all other-blaming emotions (contempt towards others and indignation towards others) for the other-agency-related action tendencies: feeling like creating a distance from one’s friend or attacking one’s friend.

## Results

### Proportion of trials for each action tendency and each agency

Figure 1 displays the proportion of trials for which each action tendency (including no-action) was selected. Means and standard deviations are presented in Supplementary Table 2. Over both groups clear differences in action tendency selection were seen between the self-agency and other-agency conditions, as expected. Feeling like attacking (self- or other-) was highly agency-specific with agency-incongruent options (e.g. feeling like attacking other in the self-agency condition) occurring rarely. In self-agency trials, apologising, creating a distance from self and attacking self were more common. In the other-agency condition, distancing from one’s friend and feeling like attacking one’s friend were more common. In contrast to feeling like reating a distance from oneself, feeling like hiding did not differ between conditions.

**Figure 1.**
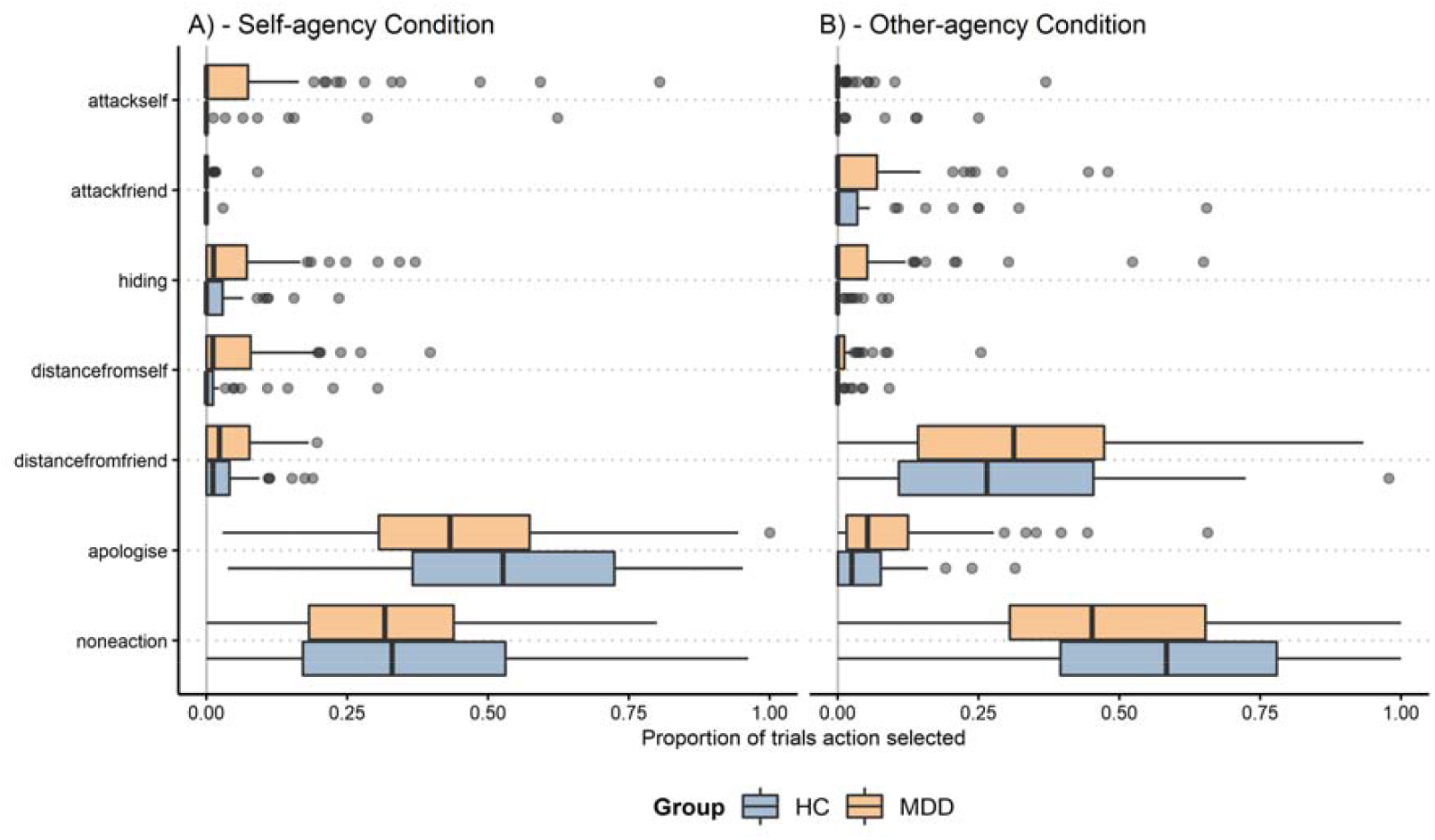
Proportion of trials action tendencies selected in both groups (HC: healthy control group; MDD: major depressive disorder group) for self-agency condition (A) and other-agency condition (B) with 95% confidence intervals as measured by the novel action tendency task.

### Group differences for choosing different action tendencies

We observed group differences that were action tendency- and agency-specific (see Table 2). Post-hoc tests for between-group differences in each action tendency over both conditions are presented in Table 3. Our first hypothesis was confirmed that MDD patients more frequently felt like hiding than control participants in both conditions and that there was a significantly higher proportion of feeling like creating a distance from oneself in the self-agency condition. In addition, there was a lower proportion of feeling like apologizing for the MDD group compared with the control group in the self-agency condition. In contrast, participants with MDD were more likely to feel like apologising or taking no action in the other-agency condition (see Table 6 for details). Participants with MDD had a significantly higher perceived control in the other-agency conditions relative to HC participants which drove the group difference in the measure of overgeneralised perception of control which we defined as the difference score between control in the self- and other-agency condition (Figure 2).

**Table 2.** Action tendencies by group and condition.

| Effect | df | F-value | p-value |
| --- | --- | --- | --- |
| group | 1, 118 | 8.95 | .003* |
| action tendency | 2.7, 323.3 | 206.45 | <.0001* |
| group x action tendency | 2.7, 323.3 | 2.59 | .06 |
| agency | 1, 118 | 0.10 | .75 |
| group x agency | 1, 118 | 5.35 | .02* |
| action tendency x agency | 3.7, 435.4 | 189.45 | <.0001* |
| group x action tendency x agency | 3.7, 435.4 | 4.16 | .003* |

The proportion of trials for which a particular action tendency was selected was arcsine square root transformed for this repeated-measures analysis of variance (ANOVA). Repeated measures ANOVA revealed a significant omnibus interaction: action tendency x agency x clinical group, $F(3.69, 435.37)=4.16$ , $p=.003$ .

**Table 3.**
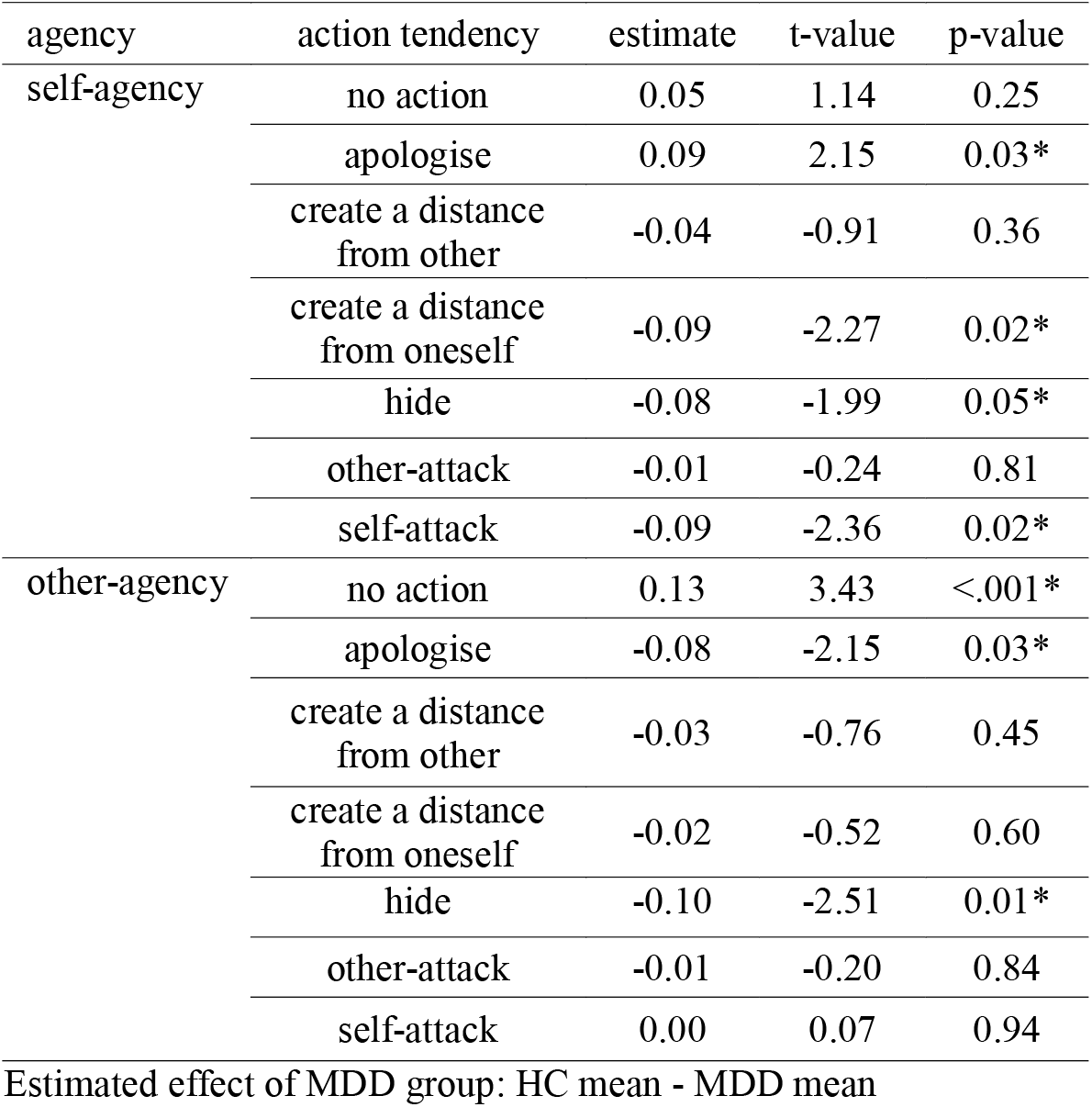
Action tendencies by group and condition - post-hoc tests.

**Figure 2.**
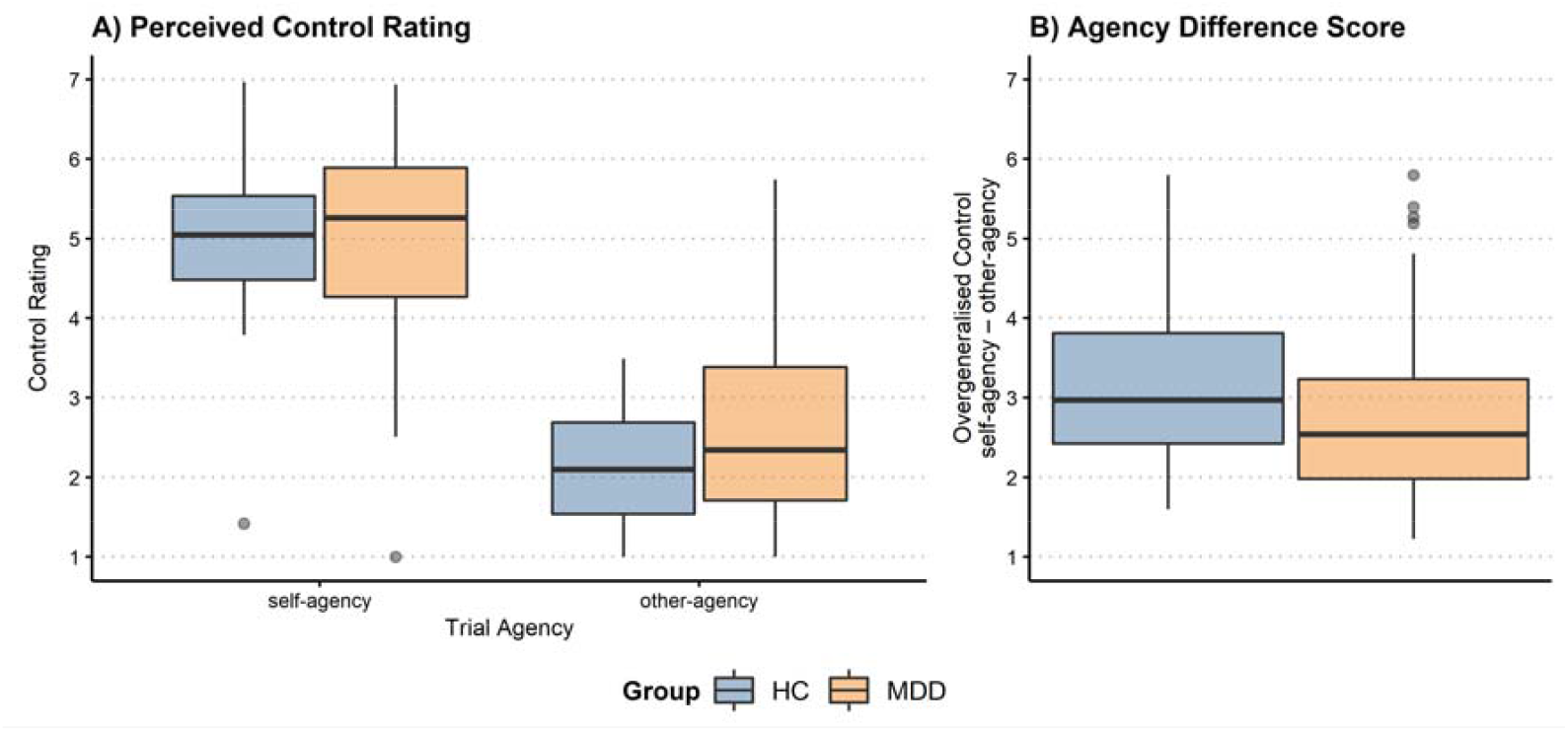
Perception of control (A) and the level of overgeneralised control (B) by clinical groups with 95% confidence intervals as measured by the valued-based moral sentiment task. Overgeneralised control was calculated by the other-agency control rating subtracted from the self-agency control rating for each subject. A higher value indicates less overgeneralisation of control ratings.

### The relationship between blame-related emotions and agency-congruent action tendencies

All self-blaming emotions were associated with a higher probability of apologising across groups (Figure 3, Tables 4 and 5). Shame and guilt were both associated with a higher probability of hiding. Interestingly, self-directed anger rather than self-disgust/contempt was associated with a higher probability of creating a distance from oneself across groups. Reversely self-disgust/contempt correlated with a higher probability of feeling like attacking oneself rather than creating a distance from oneself as we would have expected. No other main effect of group or group by emotion interactions were found for any of the other self-blame-related action tendencies (apologising, hiding, attacking oneself).

**Figure 3.**
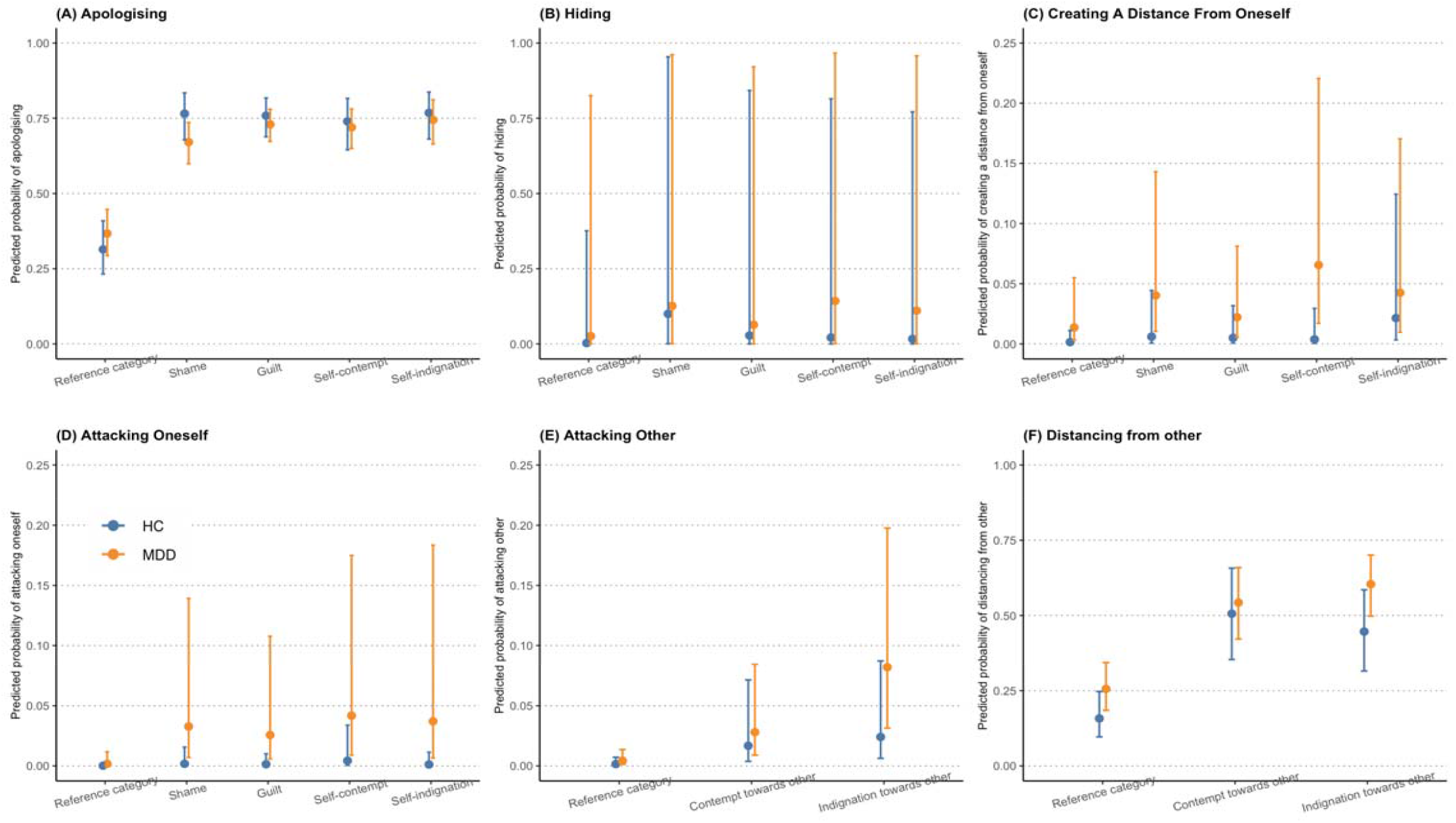
Displayed are the means and 95% confidence intervals for the predicted probability of feeling like apologising (A), hiding (B), creating a distance from oneself (C), attacking oneself (D), attacking other (E) and distancing from other (F) and their relationships to different moral emotions in both groups. (HC=Healthy Control. MDD = major depressive disorder). Reference category included trials in which participants selected other or no emotion.

**Table 4.** Self-blame-related action tendencies and emotions.

|  | Apologising vs.<br>Reference category |  |  | Hiding vs.<br>Reference category |  |  | Distancing from self vs.<br>Reference category |  |  | Attack self vs.<br>Reference category |  |  |
| --- | --- | --- | --- | --- | --- | --- | --- | --- | --- | --- | --- | --- |
|  | B | SE | p | B | SE | p | B | SE | p | B | SE | p |
| Shame | 1.96 | .22 | <.001** | 3.67 | .84 | <.001** | 1.38 | .77 | .07 | 2.31 | 1.18 | .05 |
| Guilt | 1.93 | .18 | <.001** | 2.32 | .78 | .002* | 1.20 | .63 | .06 | 2.06 | 1.10 | .06 |
| Self-disgust/contempt | 1.83 | .23 | <.001** | 2.03 | .91 | .03 | .92 | .70 | .19 | 3.20 | 1.17 | .006* |
| Self-anger | 2.00 | .23 | <.001** | 1.33 | .75 | .03 | 2.67 | .68 | <.001** | 1.89 | .152 | .13 |
| Group | .24 | .27 | .38 | 1.63 | .85 | .02 | 2.21 | 1.03 | .03 | 2.29 | 1.49 | .12 |
| Group*Shame | -.71 | .28 | .01 | -1.38 | .81 | .03 | -.27 | .88 | .76 | .65 | 1.36 | .63 |
| Group*Guilt | -.40 | .23 | .09 | -.80 | .77 | .11 | -.70 | .75 | .34 | .65 | 1.27 | .61 |
| Group*Self-disgust/contempt | -.34 | .30 | .24 | .03 | .89 | .84 | -.71 | .82 | .39 | .01 | 1.35 | .99 |
| Group*Self-Anger | -.37 | .31 | .23 | .005 | .89 | .79 | -1.50 | .87 | .08 | 1.20 | 1.47 | .41 |

Four mixed effect logistic regression models were conducted, one for each self-blame-related action tendency as outcome variable. Predictors were agency-congruent moral emotions (Shame, guilt, self-disgust/contempt, self-anger), group (MDD vs. HC) as well as their interactions as predictors. Reference categories for moral emotions in all models were the trials in which participants chose no/other emotion. The significance threshold was set to an approximate Bonferroni-corrected $p < .05$ across all 6 models (See Table 4), corresponding to an uncorrected $p < .008$ in each model. \*\*=.001, \*=.008. B=estimate, SE=standard error.

**Table 5.** Other-blame-related action tendencies and emotions.

|  | **Attacking friend vs.**<br>**Reference categories** |  |  | **Distancing from friend vs.**<br>**Reference categories** |  |  |
| --- | --- | --- | --- | --- | --- | --- |
|  | **β** | **SE** | **p** | **β** | **SE** | **p** |
| Disgust/Contempt towards friend | 2.39 | .58 | <.001** | 1.70 | .20 | <.001** |
| Anger/Indignation towards friend | 2.76 | .46 | <.001** | 1.46 | .14 | <.001** |
| Group | 1.03 | .86 | .23 | .61 | .36 | .09 |
| Group *<br>Disgust/contempt towards friend | -.50 | .73 | .49 | -.46 | .26 | .08 |
| Group *<br>Anger/indignation towards friend | .26 | .58 | .65 | .03 | .19 | .87 |

Table 6 shows that the other-blaming emotions including contempt/disgust and anger/indignation towards others were both associated with a higher probability of attacking others and distancing from others across both groups. No main effect of group nor interaction between group and other-blaming emotions were found.

**Table 6.** Perceived control ratings.

| Perceived control rating | HC (n=44) |  | MDD (n=76) |  | Welch's |  |
| --- | --- | --- | --- | --- | --- | --- |
|  | mean | SD | mean | SD | T-value | p-value |
| self-agency | 5.07 | 0.98 | 5.02 | 1.21 | .04 | .8 |
| other-agency | 2.10 | 0.72 | 2.62 | 1.16 | -.52 | .003* |
| Difference: self- vs. other-agency | 2.96 | 1.13 | 2.40 | 1.27 | .56 | .01* |

SD=standard deviation. MDD=major depressive disorder. HC=Healthy control.

### Relationship of clinical variables with maladaptive action tendencies in MDD

There were no significant correlations between proneness to feeling like hiding, or creating a distance from oneself and the number of previous depressive episodes (τ=-.13, p=.15, z=-1.46; τ=.07, p=.42, z=-.80). BDI scores were weakly positively correlated with proneness to feeling like creating a distance from oneself (τ=.31, p<.001, z=3.47), but not with feeling like hiding (τ=.14, p=.12, z=1.56).

## Discussion

We hypothesized that individuals with MDD were more likely to experience maladaptive self-blame-related action tendencies which interfere with reparative actions. We confirmed our first specific hypothesis that individuals with MDD would display an increased tendency to feel like creating a distance from themselves and like hiding. Our third specific hypothesis was supported by finding that shame was associated with feeling like hiding, in line with what has been proposed in the literature (Roseman et al., 1994; Tangney et al., 2007). However, contrary to our second hypothesis, self-contempt/disgust was associated with attacking oneself rather than feeling like creating a distance from oneself and the reverse was true of self-directed anger, further showing that emotion labels do not allow inferring predictable relationships with specific action tendencies. Intriguing was the finding of an overgeneralised perception of control for other people’s actions and a tendency to apologise for others’ wrongdoings in the MDD group.

Previous research has demonstrated the distinction between adaptive and maladaptive action tendencies. The former promotes constructive and proactive pursuit, such as apologizing, whereas the latter motivates defensiveness, social withdrawal and interpersonal separation, such as hiding and creating a distance from oneself (Tangney et al., 2007). Given a lack empirical studies, our study is the first to demonstrate that maladaptive action tendencies distinguish participants with MDD and healthy control participants, who were closely matched apart from their difference in MDD vulnerability. Maladaptive action tendencies as demonstrated here could contribute to maladaptive coping styles, such as avoidance-oriented coping, which predicted anxious and fearful responding under stressful circumstances (Spira, Zvolensky, Eifert, & Feldner, 2004) and have been consistently found to be associated with MDD (Berghuis & Stanton, 2002; Burker, Evon, Losielle, Finkel, & Mill, 2005). As such, creating a distance from oneself and hiding might motivate people to use social avoidance as a coping mechanism, which could contribute to their MDD vulnerability in response to negative social feedback. From a more theoretical perspective, feeling like creating a distance from oneself might also involve the rejection and denial of one’s self-identity. This could be a purer measure than self-disgust or self-contempt of a maladaptive discrepancy between the actual self and the ought self, proposed by Higgins (1987). An increase in self-discrepancy was both directly and indirectly linked to depressive symptoms (Roelofs et al., 2007) and possibly also related to one’s vulnerability to MDD.

In addition, the finding that people with MDD tend to attribute control to themselves and apologise more readily when their friend has done something wrong could be explained by our previous finding that individuals with MDD have increased overgeneralised self-blaming emotions and reduced blame-related emotions towards others (Zahn, Lythe, Gethin, Green, Deakin, Workman, et al., 2015). This demonstrates that overgeneralised control over other people’s wrongdoing could be also a vulnerability trait of MDD which is in keeping with increased omnipotent responsibility guilt in MDD (O’Connor et al., 2002).

On the other hand, our findings are inconsistent with the view that shame is specifically associated with maladaptive action tendencies that promote defensiveness, interpersonal separation and distance, and only guilt is specifically associated with adaptive action tendencies that motivate constructive and reparative actions such as apologising (Ketelaar & Tung Au, 2003; Tangney et al., 2007). Instead, our results show that all self-blaming emotions, including shame, guilt, self-contempt/disgust and self-indignation were associated with a tendency to apologise, but that guilt and shame can also both be associated with feeling like hiding. These differences could be partially explained by different measures used to assess blame-related emotions. For example, the Test of Self-Conscious Affect-3 (TOSCA-3) developed by Tangney J. P., Dearing R. L., Wagner P. E., and R. (2000) was different from our measure of self-blame where participants were directly asked to label their moral emotions. The former has been suggested to only assess the adaptive form of guilt and the maladaptive form of shame (Luyten, Fontaine, & Corveleyn, 2002). Other researchers suggested that the TOSCA measured motivation rather than moral emotions (Giner-Sorolla, Piazza, & Espinosa, 2011). Indeed, the various methods used in the literature to measure blame-related emotions made it difficult to compare findings across studies. As such, a more unified and standardized measure of self-blame that addresses both adaptive and maladaptive forms of blame-related emotions is important in future studies.

### Limitations

On a more cautionary note, our study was limited firstly by its cross-sectional design, which made it difficult to infer a causal relationship between MDD vulnerability and action tendencies. While maladaptive action tendencies could be a vulnerability trait for MDD, it is also possible that these represent scarring effects of previous depressive episodes (Wichers, Geschwind, Van Os, & Peeters, 2010). Nevertheless, this is unlikely as no correlation was found between maladaptive action tendencies and the number of previous depressive episodes. Future studies are needed to determine whether feeling like creating a distance from oneself is a pure trait marker of vulnerability or is also modulated by depressive state which its correlation with residual symptoms suggests. Secondly, the task was in a verbal format and included abstract descriptions of scenarios. Thus, participants’ self-blaming emotions might have depended in part on how well they can imagine the scenarios and indeed we previously found an association between visual imagery ratings and emotional intensity ratings (Zahn et al., 2009). Furthermore, we have previously found that structural anatomical differences in posterior cortical areas known to be relevant for visual imagery could partly explain individual differences on the VMST (Zahn, Garrido, Moll, & Grafman, 2014). It is therefore important to develop more immersive tasks to measure blame-related action tendencies in future studies, which rely less on the ability to create one’s own imagery, which recent research shows is a widely varying ability (Fulford et al., 2018).

### Conclusions

Taken together, feeling like creating a distance from oneself and hiding were distinctive for remitted MDD compared with the control group, thus unveiling a novel marker of psychopathology, which was present even when symptoms had subsided. Future studies are needed to probe the prognostic value of maladaptive action tendencies. If replicated, our findings further suggest the development of novel psychological and neurocognitive treatments specifically aiming at self-distancing and hiding which are so far neither assessed nor addressed in standard psychotherapeutic approaches.

## Data Availability

Data is available from the corresponding author upon reasonable request.

## Acknowledgements

We thank all the participants who have taken part in our study. We are grateful to Dr Karen Lythe for her important contribution to design and data collection. We are also grateful to Dr Jennifer Gethin and Dr Clifford Workman for their important contribution to recruitment and assessment of participants. We further thank Professors Bill Deakin and Matthew Lambon-Ralph for their important contributions as co-investigators of the project.

