## supplemental tables for "The role of blame-related action tendencies in the vulnerability to major depressive disorder"

**Supplementary Material**

**Supplementary Table 1 | Exclusion reasons for participants following phone pre-screening interview.**

| Reason for exclusion | n |
| --- | --- |
| Current antihypertensive medications or statins | 20 |
| Current antidepressant or other centrally active medications | 52 |
| Diabetes | 4 |
| Epilepsy | 5 |
| Multiple sclerosis | 3 |
| Past cancer | 7 |
| Past stroke | 1 |
| Thyroid function problems | 19 |
| Vitamin D deficiency | 1 |
| Other psychiatric disorders than MDD | 54 |
| Substance or alcohol abuse | 23 |
| Other general medical condition | 5 |
| Family history of MDD/bipolar/schizophrenia (control group) | 26 |
| Excluded because of age-matching (control group) | 3 |
| Left-handed | 20 |
| MRI contraindications | 77 |
| Non-native English speaker | 19 |
| Out of age range | 4 |
| No reason recorded | 5 |
| Withdrawal after phone pre-screening | 33 |
| Not meeting full screening criteria for MDD | 30 |
| Not remitted for long enough | 7 |
| Fulfilling criteria for current MDD | 13 |
| Total excluded after phone pre-screening | 431 |

**Supplemental Table 2| Means and standard deviations of proportion of trials for which a particular action tendency was chosen**

| Action | Healthy Control (n=44) | | MDD (n=76) | | |
| --- | --- | --- | --- | --- | --- |
|  | self-agency | other -agency | | self-agency | other-agency |
| No action | 0.356 (0.233) | 0.572 (0.282) | | 0.320 (0.206) | 0.467 (0.257) |
| Apologise | 0.530 (0.230) | 0.050 (0.068) | | 0.451 (0.219) | 0.093 (0.120) |
| Distance from  friend | 0.034 (0.051) | 0.296 (0.234) | | 0.046 (0.055) | 0.319 (0.230) |
| Distance from self | 0.023 (0.061) | 0.006 (0.017) | | 0.055 (0.081) | 0.012 (0.033) |
| Hide | 0.025 (0.050) | 0.008 (0.020) | | 0.056 (0.085) | 0.049 (0.107) |
| Attack friend | 0.001 (0.004) | 0.053 (0.121) | | 0.002 (0.011) | 0.051 (0.095) |
| Attack self | 0.032 (0.106) | 0.015 (0.048) | | 0.070 (0.143) | 0.011 (0.045) |
